# Prevalence of Internet Usage Complaints Among Undergraduate Students

**DOI:** 10.1101/2024.01.29.24301866

**Authors:** Mgbedo Nnaemeka Emmanuel, Nana Zavradashvili

## Abstract

Internet access has aided lots of students in achieving academic purposes. This study aimed to examine the prevalence of internet usage complaints on students health, during mandatory online studies among undergraduate students from the University of Georgia (UG), Tbilisi. This cross-sectional was conducted among undergraduate students from UG using a web designed questionnaire. The survey contained sociodemographic characteristics, internet access, and health complains. The study was reviewed and approved by the Institutional Review Board (IRB). Most of participants were females (70.5%), from School of Health Sciences (207, 32.9%), in first year of study (354, 56.2%), and usually spend time on social media (Facebook, Instagram, Twitter, WhatsApp, etc.) (315, 50%). Students reported different complaints such as internet craving, feeling guilty after internet use, internet dyscontrol, back, and neck pain. There was no significant difference in sociodemographic variables. Our findings suggested that the prolonged use of internet could have some health implications among students.

## Introduction

The widespread of the popularity of the viral disease Covid-19 [1] led to online studies that could have increased access to the Internet with an impact on students’ habits towards their academics [2]. Despite the surge in the number of cases, there were changes in the level of physical and social activities, especially among students [3, 4] which could have increased their access to the Internet. The rate of Internet accessibility worldwide has increased as it is used to achieve a desired task [5]. According to the National Statistics Office of Georgia, the rate of frequent Internet users has increased from 89.2% in 2019 to 92.8% in 2022 [6]. Access has helped students to acquire knowledge and improve academic performance [7] and research skills [8]. It can be used to satisfy social and entertainment drives such as listening to online music, watching movies, and playing video games [9–12]. The Internet has been considered a stress-coping mechanism [13, 14] while watching online programs or gaming but can lead to addiction [15]. Students tend to spend more time on the Internet, which can affect their physical health, resulting in pain in the eyes, fingers, neck, and shoulder, and clinical symptoms such as dry eyes, headaches, vision problems, and loss of appetite [16-18]. This study aimed to examine the prevalence of internet usage complaints on students’ health, during mandatory online studies among undergraduate students from the University of Georgia (UG), Tbilisi.

## Materials and Methods

### Study Population and Sampling

The study collected 630 responses from undergraduate students studying in a different department at the University of Georgia during the online academic session. All questionnaires were web-based, as each participant was informed of the purpose of the study. The study was voluntary and anonymous, as no reward was offered to the participants. The participants were asked to respond only once, as the use of the same IP address was restricted to ensure validity of the study.

### Data Collection

Data were collected using self-administered web-based questionnaires shared with the students through the university intranet after receiving ethical approval from the university board. The survey lasted for one week, and the link was opened from November 10th to 18th, 2022.

### Demographic variables

Demographic variables included demographic characteristics (such as gender, age (indicated by students in years), undergraduate program, year of study, religion, and frequent activity performed.

### Dependent Variable

The level of physical activity was assessed as the participants were asked if they had an average sleep duration of more than seven (7) hours in the past week. The students’ mental health was assessed using five (5) items. A closed-ended response (yes or no) was required, as the questions were related to Internet craving, feelings, tolerance, withdrawal, and dyscontrol.

The seven (7) physical complaints reported were used to assess individual physical health to determine if they had suffered from internet-associated problems during or after use. Such complaints included back pain, finger numbness, neck pain, headache, inability to sleep, vision problems, and loss of appetite, which were computed scored as yes or no.

### Principal Study Variable

The principal study variables were individual access to the internet and gender. Daily Internet accessibility was categorized as seldom, casual, regular, frequent, or intense. The categories were defined as seldom (less than an hour), casual (between 1-3 hours per day), regular (between 3 hours-5 hours), frequent (between 5 hours-7 hours), and intense (above 7 hours).

## Statistical Analysis

The statistical analyses were performed using the Statistical Package for the Social Sciences (SPSS) version 23.0 software (SPSS Inc., Chicago, IL, USA) mainly the descriptive analysis; a t-test was used to compare means of more than two independent groups and statistically significant at p<0.05. Physical and mental health responses (yes/no) were categorized as dichotomous responses before performing the linear regression. Linear regression was performed to check for multicollinearity, homogeneity of variance, and Variance Inflation Factors (VIF <4). Prevalence was analyzed using standard error at a 95% confidence interval (95% CI).

## Results

Table 1 shows demographic characteristics such as gender, age, discipline, year of study, religion, work status, physical activity, weekly average sleep duration, and frequent activity. The majority of participants were female (444, 70.5%), 2.7 years old in the age category of 17-21 years of age (80.2%), from the School of Health Sciences (207, 32.9%), in the first year of study (354, 56.2%), and in the Christian religion (460, 73%), not working (497, 78.9%), sometimes performed physical activity sometimes (415, 65.9%), had an average weekly sleep of less than 7 hours (326, 51.7%), and usually spent time on social media (Facebook, Instagram, Twitter, WhatsApp, etc.) (315, 50%).

**Table 1.0.**
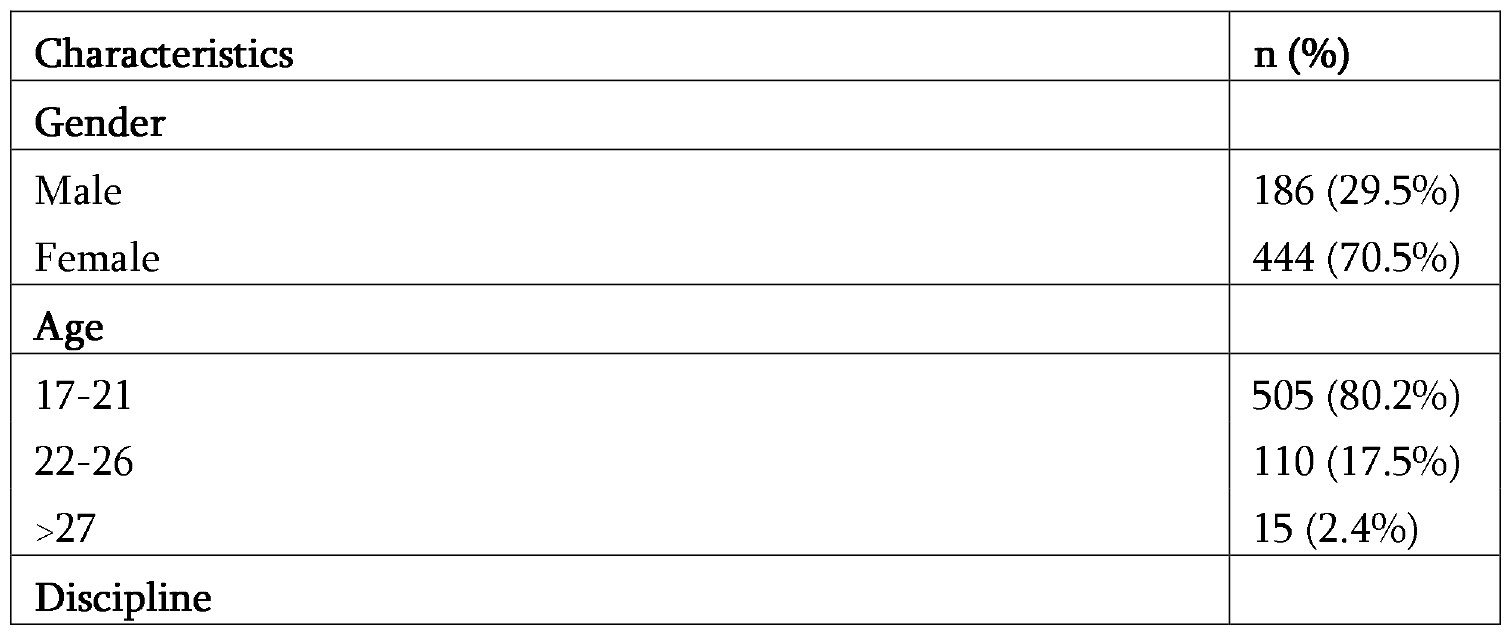
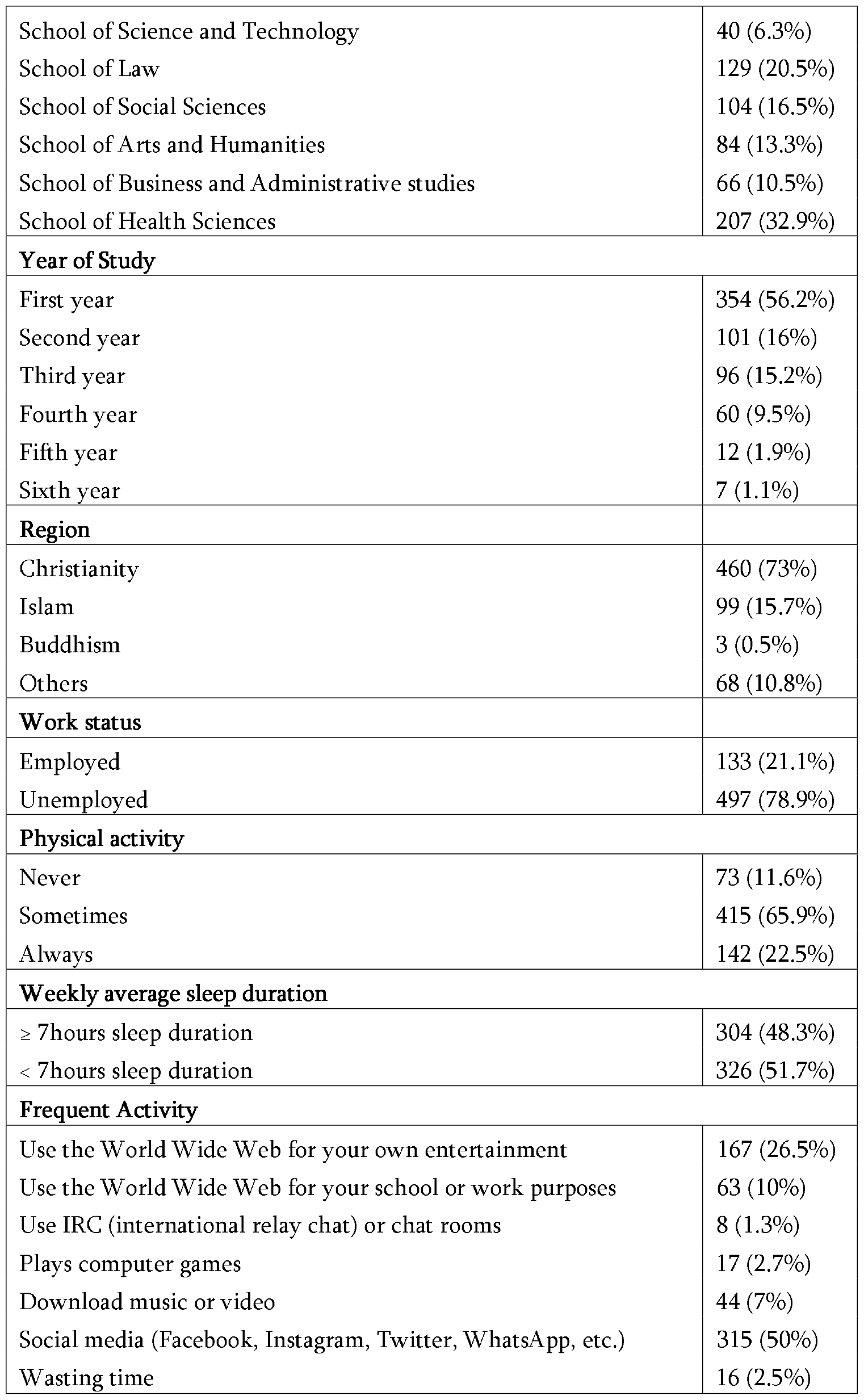
Demographic characteristics of Undergraduate students in the UG.

| Characteristics | n (%) |
| --- | --- |
| **Gender** |  |
| Male | 186 (29.5%) |
| Female | 444 (70.5%) |
| **Age** |  |
| 17-21 | 505 (80.2%) |
| 22-26 | 110 (17.5%) |
| >27 | 15 (2.4%) |
| **Discipline** |  |
| School of Science and Technology | 40 (6.3%) |
| School of Law | 129 (20.5%) |
| School of Social Sciences | 104 (16.5%) |
| School of Arts and Humanities | 84 (13.3%) |
| School of Business and Administrative studies | 66 (10.5%) |
| School of Health Sciences | 207 (32.9%) |
| **Year of Study** |  |
| First year | 354 (56.2%) |
| Second year | 101 (16%) |
| Third year | 96 (15.2%) |
| Fourth year | 60 (9.5%) |
| Fifth year | 12 (1.9%) |
| Sixth year | 7 (1.1%) |
| **Region** |  |
| Christianity | 460 (73%) |
| Islam | 99 (15.7%) |
| Buddhism | 3 (0.5%) |
| Others | 68 (10.8%) |
| **Work status** |  |
| Employed | 133 (21.1%) |
| Unemployed | 497 (78.9%) |
| **Physical activity** |  |
| Never | 73 (11.6%) |
| Sometimes | 415 (65.9%) |
| Always | 142 (22.5%) |
| **Weekly average sleep duration** |  |
| ≥ 7 hours sleep duration | 304 (48.3%) |
| < 7 hours sleep duration | 326 (51.7%) |
| **Frequent Activity** |  |
| Use the World Wide Web for your own entertainment | 167 (26.5%) |
| Use the World Wide Web for your school or work purposes | 63 (10%) |
| Use IRC (international relay chat) or chat rooms | 8 (1.3%) |
| Plays computer games | 17 (2.7%) |
| Download music or video | 44 (7%) |
| Social media (Facebook, Instagram, Twitter, WhatsApp, etc.) | 315 (50%) |
| Wasting time | 16 (2.5%) |

As shown in table 2, most of the students who reported intense daily use of the Internet had some mental health reports on craving (298, 56.4%), feeling (198, 57.9%), tolerance (208, 59.8%), withdrawal (195, 62.3%), and dyscontrol (245, 7.2%).

**Table 2.0.**
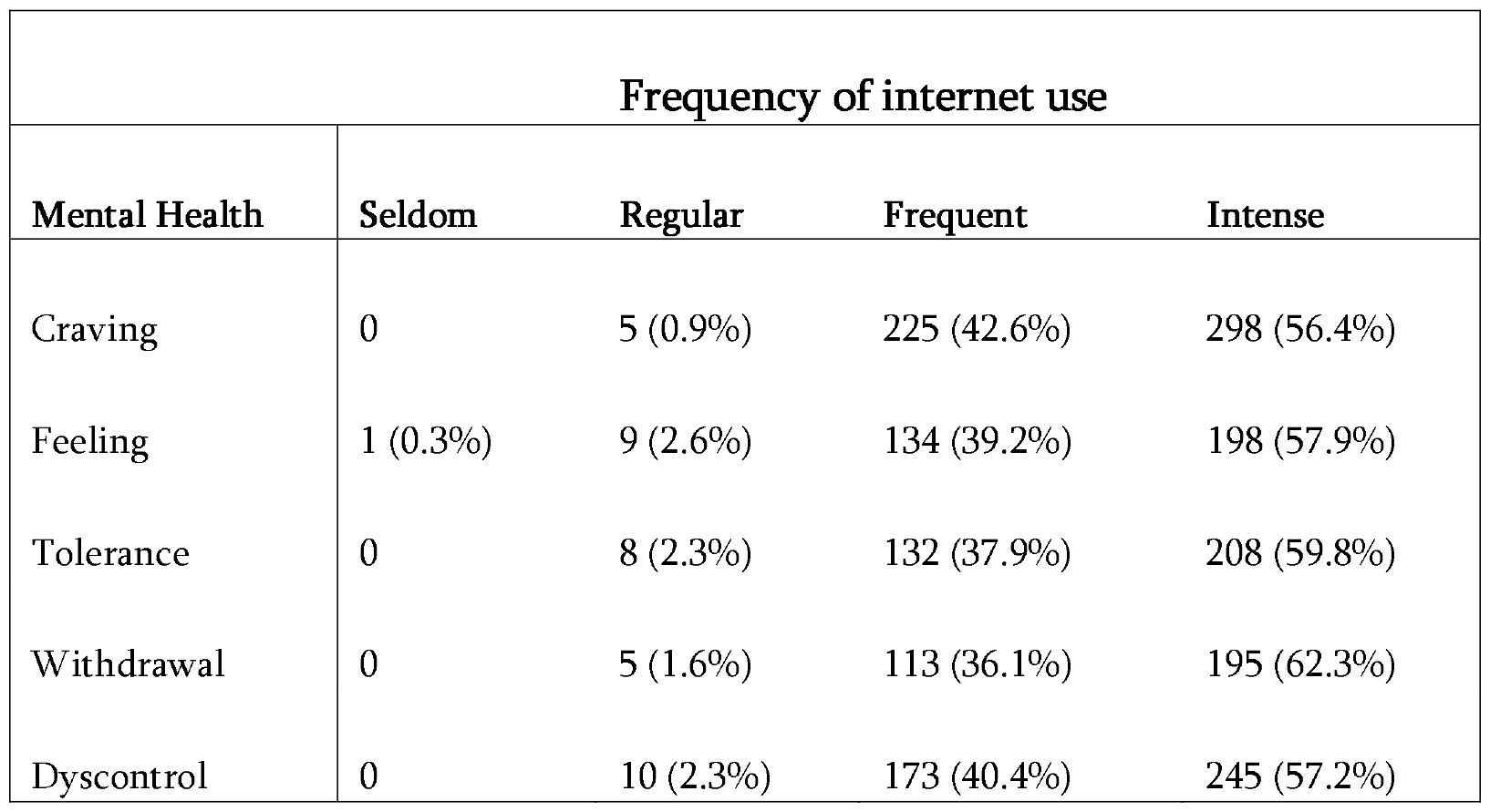
Demographic characteristics according to the frequency of internet use and mental health among the students.

In Table 3, most of the students who intensely use the internet daily indicated certain complaints including, Inability to sleep (148, 57.4%), headache (147, 53.5%), vision problems (121, 54.3%), back pain (102, 55.4%), neck pain (99, 56.6%), finger numbness (47, 58%), and the least of the complaint was the loss of appetite (32, 53.3%).

**Table 3.0.** Demographic characteristics according to the frequent internet use and physical complaints among students.

| Physical Complaints | Frequency of internet use |  |  |  |  |
| --- | --- | --- | --- | --- | --- |
|  | Seldom | Casual | Regular | Frequent | Intense |
| Back Pain | 0 | 0 | 8 (4.3%) | 74 (40.2%) | 102 (55.4%) |
| Finger numbness | 0 | 1 (1.2%) | 2 (2.5%) | 31 (38.3%) | 47 (58%) |
| Neck pain | 0 | 0 | 6 (3.4%) | 70 (40%) | 99 (56.6%) |
| Headache | 1 (0.4%) | 0 | 9 (3.3%) | 118 (42.9%) | 147 (53.5%) |
| Inability to sleep | 1 (0.4%) | 0 | 5 (1.9%) | 104 (40.3%) | 148 (57.4%) |
| Vision Problems | 1 (0.4%) | 0 | 7 (3.1%) | 94 (42.2%) | 121 (54.3%) |
| Loss of appetite | 0 | 0 | 2 (3.3%) | 26 (43.3%) | 32 (53.3%) |
| No complaint | 0 | 0 | 0 | 5 (45.5%) | 6 (54.5%) |

Table 4 presents the frequency of mental health, physical complaints, and risks associated with females. The results displayed the differences according to gender, in which the female students were at high risk for Internet craving (OR 0.536, 95% CI 0.346-0.83), guilty or depression after using the Internet (feeling) (OR 0.651, 95% CI 0.462-0.919), and Internet dyscontrol (OR 0.537, 95% CI 0.375-0.767). Moreover, the female students were at risk of having certain physical health complaints, such as back pain (OR 1.629, 95% CI 1.13-2.349) and neck pain (OR 2.018, 95% CI 1.396-2.918).

**Table 4.0.** Comparison of the students’ mental health, physical complaints, and Odds Ratio (OR) depending on gender.

|  |  |  |  |  |  |  |  | 95% Confidence Interval (CI) |  |
| --- | --- | --- | --- | --- | --- | --- | --- | --- | --- |
| Mental Health |  | Male | Female | Total | Chi-square | p-Value | Odd Ratio | Lower | Upper |
| Craving | Yes | 144 (77.4%) | 384 (86.5%) | 528 (83.8%) | 7.942a | **0.005** | 0.536 | 0.346 | 0.83 |
|  | No | 42 (22.6%) | 60 (13.5%) | 102 (16.2%) |  |  |  |  |  |
| Feeling | Yes | 87 (46.8%) | 255 (57.4%) | 342 (54.3%) | 6.001a | **0.014** | 0.651 | 0.462 | 0.919 |
|  | No | 99 (53.2%) | 189 (42.6%) | 288 (45.7%) |  |  |  |  |  |
| Tolerance | Yes | 103 (55.4%) | 245 (55.2%) | 348 (55.2%) | .002a | 0.964 | 1.008 | 0.714 | 1.422 |
|  | No | 83 (44.6%) | 199 (44.8%) | 282 (44.8%) |  |  |  |  |  |
| Withdrawal | Yes | 89 (47.8%) | 224 (50.5%) | 313 (49.7%) | .355a | 0.551 | 0.901 | 0.64 | 1.269 |
|  | No | 97 (52.2%) | 220 (49.5%) | 317 (50.3%) |  |  |  |  |  |
| Dyscontrol | Yes | 108 (58.1%) | 320 (72.1%) | 428 (67.9%) | 11.808a | **0.001** | 0.537 | 0.375 | 0.767 |
|  | No | 78 (41.9%) | 124 (27.9%) | 202 (32.1%) |  |  |  |  |  |
| Physical complaints |  |  |  |  |  |  |  |  |  |
| Back pain | Yes | 68 (36.6%) | 116 (26.1%) | 184 (29.2%) | 6.901a | **0.009** | 1.629 | 1.13 | 2.349 |
|  | No | 118 (63.4%) | 328 (73.9%) | 446 (70.8%) |  |  |  |  |  |
| Finger numbness | Yes | 22 (11.8%) | 59 (13.3%) | 81 (12.9%) | 0.250a | 0.617 | 0.875 | 0.519 | 1.476 |
|  | No | 164 (88.2%) | 385 (86.7%) | 549 (87.1%) |  |  |  |  |  |
| Neck pain | Yes | 71 (38.2%) | 104 (23.4%) | 175 (27.8%) | **14.213a** | **0.000** | **2.018** | **1.396** | **2.918** |
|  | No | 115 (61.8%) | 340 (76.6%) | 455 (72.2%) |  |  |  |  |  |
| Headache | Yes | 88 (47.3%) | 187 (42.1%) | 275 (43.7%) | 1.438a | 0.230 | 1.234 | 0.875 | 1.741 |
|  | No | 98 (52.7%) | 257 (57.9%) | 355 (56.3%) |  |  |  |  |  |
| Inability to sleep | Yes | 80 (43%) | 178 (40.1%) | 258 (41%) | .462a | 0.496 | 1.128 | 0.797 | 1.595 |
|  | No | 106 (57%) | 266 (59.9%) | 372 (59%) |  |  |  |  |  |
| Vision Problems | Yes | 70 (37.6%) | 153 (34.5%) | 223 (35.4%) | .578a | 0.447 | 1.148 | 0.804 | 1.638 |
|  | No | 116 (62.4%) | 291 (65.5%) | 407 (64.6%) |  |  |  |  |  |
| Loss of appetite | Yes | 24 (12.9%) | 36 (8.1%) | 60 (9.5%) | 3.498a | 0.061 | 1.679 | 0.971 | 2.903 |
|  | No | 162 (87.1%) | 408 (91.9%) | 569 (90.3%) |  |  |  |  |  |

Table 5 displays the mean scores of health reports (mental and physical complaints) and their associations with sociodemographic variables. The mean scores varied across the predictor variables, including sex, discipline, religion, craving, feeling, tolerance, withdrawal, dyscontrol, back pain, finger numbness, neck pain, headache, inability to sleep, vision problems, and loss of appetite. Variables such as craving, tolerance, and withdrawal were significant because of the prolonged use of the internet among students.

**Table 5.0.**
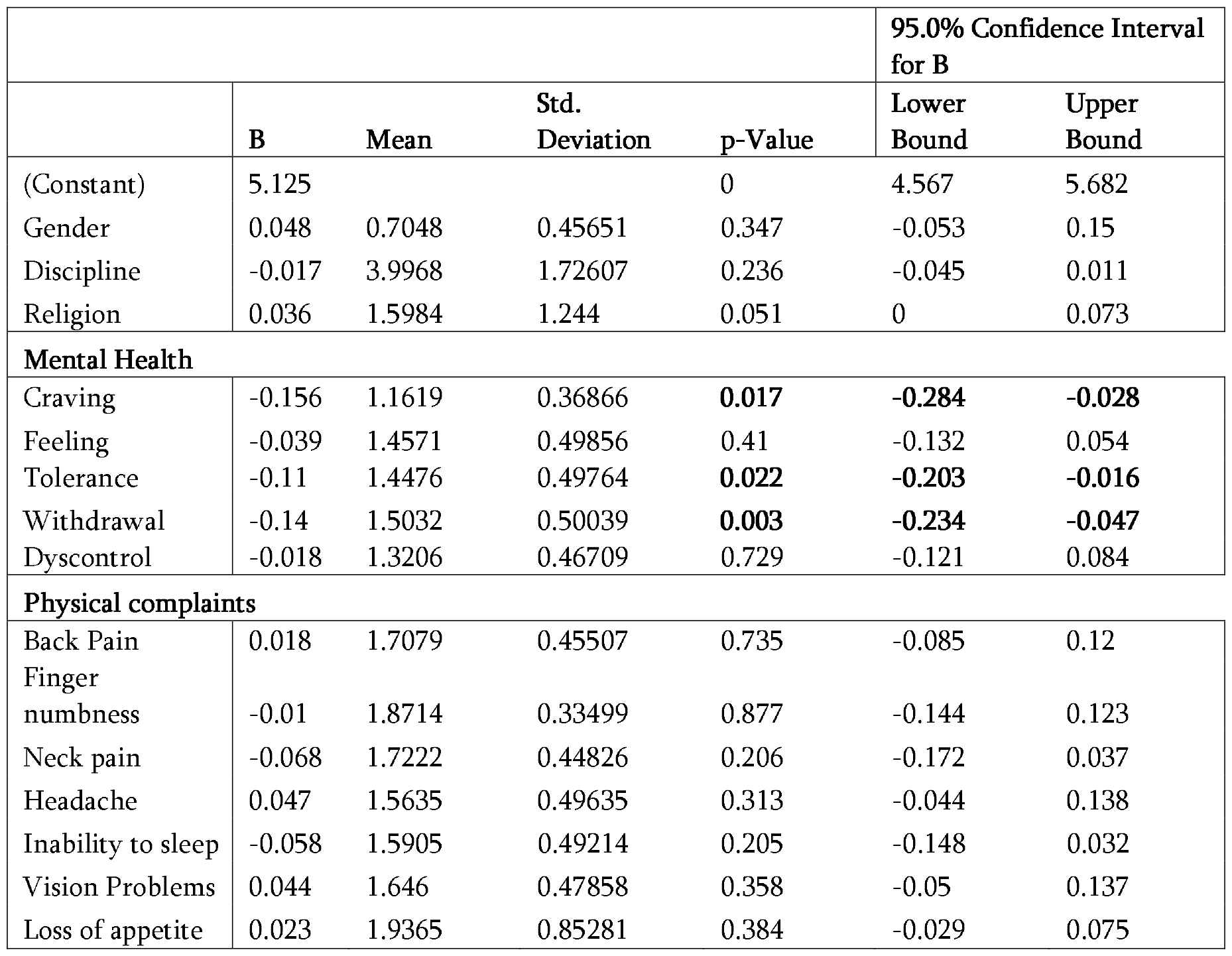
Results of simple linear regression for the mean of variables depending on internet daily access.

## Discussion

The influence of online studies during the Covid pandemic has led to different investigative research studies on Internet use, accompanied by problematic health complaints among students [16, 19]. In this study, no significant difference was found in sociodemographic variables, like the 9-month longitudinal study regarding problematic social media use, gaming, and psychological distress among Hong Kong and Taiwan University students [20]. However, the participation of the females compared to male students from different fields of study in UG, use the Internet more for different activities. Although there was an assessment for some specific activities popular among male students, such as gaming [21], the participants were low in comparison with other activities popular among the females, such as social media [22].

There are certain complaints prevailing among the frequent and intense Internet users compared with the seldom and regular users. Craving, feeling, and dyscontrol were the most perceived mental health complaints among the female students. Studies have shown that social media can be associated with certain depressive symptoms [23] which could have an impact on the students’ mental health. Overall, craving, tolerance, and withdrawal were the predominant complaints of students. Prolonged Internet use can cause psychological problems which were reported among university students in China [24].

This study observed that back and neck pain was the most prevalent symptoms among female students [25]. Studies [26] reported that the frequent and excessive use of the Internet could cause certain physical complaints among undergraduate students, such as headache, back pain, finger numbness, and neck pain; however, the complaints of back pain and neck pain were consistent in this study. Neck pain is a common symptom that has been reported over time among the students at the University of Gondar, Ethiopia [27] because of the overuse of the Internet. The use of internet continues to grow as most students’ complaints of clinical symptoms which could be associated to frequency of use. Further comprehensive studies are required to properly control the psychiatry impact which these could result in.

## Limitation

The study was conducted among the undergraduate students from a private University which restricts it generalization to all students in Georgia country. We could not determine how long students spend on each social media and the possible complaints. Despite the limitations of our study, the sample size was large. The students were required to indicate their university code for consultations regarding their complaints which validates the responses.

## Data Availability

All data produced in the present study are available upon reasonable request to the authors

## Acknowledgement

We appreciate the effort of the students who participated in this study and the University of Georgia for the study approval.

## Author Contribution Statement

The authors confirm contribution to the article as follows: study concept and design, data collection: Nnaemeka Emmanuel Mgbedo; analysis and interpretation of results, draft manuscript preparation; Nnaemeka Emmanuel Mgbedo, Nana Zavradashvili. All authors reviewed the results and approved the final version of the manuscript.

## Ethics Approval statements

The study was conducted in accordance with the Declaration of Helsinki and approved by the Institutional Review Board of the School of Health Sciences, the University of Georgia (UGREC: 11-24272)

